# Factors Influencing 4th Year Medical Students’ Rank Lists of Radiology Residency Programs

**DOI:** 10.1101/2020.06.29.20142968

**Authors:** Florentino Saenz Rios, Shadan Alwan, Quan Nguyen

## Abstract

**Background:** Applicants for radiology residency programs consider various factors when completing their ranking lists of programs. However, in making their decision, there is usually minimal objective data that can guide applicants in creating their rank list.

**Purpose:** The goal of the study is to determine which factors in order of importance lead to the decision to rank one residency program over the other.

**Methods:** This study was an anonymous, cross-sectional study conducted through an online survey. Emails were sent out to applicants who had interviewed at the University of Texas Medical Branch’s Diagnostic Radiology department the day after match day. The survey consisted of a single open-ended question asking the survey taker to list four factors in order of importance that guided their program rank list. Results were compiled, tallied, and categorized to find common themes between the applicant’s preferences.

**Results:** Of the 73 surveys sent out, a total of 30 applicants responded for a response rate of 41%. The most common “1^st^ preference” factors were “Reputation,” “Location,” and “Work Environment,” with 23.33%, 20%, and 20% of applicants considering these the most important factors in ranking a program.

**Conclusion:** Despite reputation being the most common “1^st^ preference” factor, “Work Environment” was more commonly mentioned overall, being mentioned in 23.35% of all answers compared to “Reputation” 12.5%. These results, combined with the difficulty in changing a program’s location and reputation, make it clear that programs should focus on building and highlighting a healthy work environment to attract applicants.

## Introduction

There are many factors Diagnostic Radiology residency applicants can consider during the stressful process of the NRMP residency match. While each individual may have different personal preferences for what they want out of this next monumental step, it may be possible to find common threads among applicants. Also, trends in the fluctuating competitiveness of radiology make attracting applicants that are both high quality and a good fit for the program difficult.^[1, 3–5, 8]^ In addition, it is possible that potential resident burnout could be minimized if applicants receive more exposure to factors that residents consider important, allowing a higher program-resident fit to be met by allowing applicants to find a program that most meets the factors they consider in job satisfaction.^[1, 2, 6, 8]^ Due to this, this study aims to determine what factors applicants prioritize when deciding where a program will fall in an applicant’s rank list. There is currently limited data to guide Diagnostic Radiology program directors or residency selection committees, ultimately making the process of highlighting the program’s strengths to applicants more difficult and inefficient. Programs may be misallocating their limited interview time into presenting factors that may align with that program’s strengths, but applicants may not find as appealing. ^[1, 3, 6]^ Objective data into applicant preferences may act as a catalyst for programs to implement or modify their interview strategies and curriculum to provide a higher quality education while also realigning their recruitment efforts to highlight the program’s strengths in accordance with the applicant’s preferences.

## Methods

This study was an anonymous, cross-sectional study conducted via means of an online survey. The survey was disbursed among all Allopathic and Osteopathic applicants applying to the University of Texas Medical Branch (UTMB) Diagnostic Radiology residency programs during the 2019 NRMP match season. This study received IRB approval from the IRB as well as an exemption from requiring informed consent due to the anonymous design of the study. No compensation was offered to candidates for their participation in the survey.

### Enrollment into the Survey

This study aims to determine what ranking factors Diagnostic Radiology applicants consider when creating their rank list. As such, the study population consisted of fourth-year medical students applying for a first□year position at the Diagnostic Radiology program at UTMB (Galveston, TX). Candidate’s email information was provided to UTMB from the applicant list on the Electronic Residency Application Service (ERAS, Washington, DC). In total, 73 candidates were emailed the survey the day after the 2019 NRMP match day on March 16^th^, 2019. The survey was made available for two weeks after initial contact, closing on March 30^th^, 2019. Candidates were contacted via the ERAS provided email and were informed of the purpose of the survey, a statement that the survey was independent of UTMB’s selection process, and the availability of the option to opt-in or out by responding or not responding to the survey.

### Development of the Survey

The structure of the survey was developed through a review of relevant literature ^[1, 2, 4, 5]^, discussion among the investigating authors, and a focus group with 15 volunteering UTMB Diagnostic Radiology residents of different years. To allow participants to anonymously express their priorities when determining their rank list in full, it was decided that the online survey website Survey Monkey™ (Palo Alto, Ca) would be used. It was determined that in order to both maximize survey response rates and eliminate bias from predetermined answer choices that the survey would be developed to include one mandatory open-ended. Participants who chose to take the survey were instructed to “Describe the top four factors you consider from most to least important when ranking a Diagnostic Radiology program?” Upon completion of the survey, participants were thanked for their participation in addition to being reminded that the survey was independent of the participating program’s selection process. The responses to the open-ended question were compiled, grouped into categories, and tallied amid discussion by the investigating authors and volunteering UTMB residents.

### Disbursement of the Survey

Potential participants were emailed all information on participation in the study, including study purpose, reminders, and survey link, through the blind “carbon copy” feature to provide participants anonymity from one another. Email invitations were sent to each potential participant on March 16^th^, 2019, the day after the NRMP match. In this email, potential participants were once again informed that participation was not mandatory and that the survey would remain open for the following two weeks from March 16^th^, 2019 to March 30^th^, 2019. Completed surveys that were submitted before the closure of the survey on March 30^th^, 2019, were eligible for data analysis.

## Results

A total of 73 emails (n = 73) were gathered from ERAS with 73 invitations to participate in the study sent out. Of these 73, a total of 30 participants completed the survey, giving the survey a response rate of approximately 41%. The answers to each of the 30 surveys completed were compiled, grouped into categories, and tallied amid discussion by the investigating authors and volunteering UTMB residents. After grouping of the participants’ answers, the factors included “Location,” “Reputation,” “Work Environment,” “Education,” “Work-Life Balance,” “Fellowships,” “Board Pass Rate,” “Moonlighting,” “Program Size,” “Faculty: Resident Ratio,” “Family & Friends,” “Case Volume/Diversity,” “Research,” and “Other.” The factor “Other” was reserved for any answers that did not fit within any of the themes of the other factors decided on. Due to no underlying theme that residency programs can use as a means to improve, the “Other” factor will not be discussed in this analysis.

### Factors Considered The 1^st^ Preference

As seen in figure 1, survey findings found that the single most mentioned factor influencing a program’s placement in the rank list was a program’s “Reputation,” with approximately 23.33% of surveyed MS4s listing it as their “1^st^ preference” ranking factor. This was closely followed by both “Work Environment” and “Location,” which were both mentioned in approximately 20% of the applicant’s answers as the second most mentioned ranking factor. Factors such as “Education,” “Family & Friends,” “Case Volume/Diversity” all came tied for the third-most mentioned “1^st^ preference” factor, with each of these factors being mentioned by approximately 6.67% of applicants surveyed. Both of the ranking factors “Board Pass Rate” and “Research” were only mentioned as an applicant’s “1^st^ preference” rank factor by 3.33% of all applicants surveyed. The ranking factors “Fellowship,” “Moonlighting,” “Faculty: Resident Ratio,” “Program Size,” and “Work-Life Balance” were not mentioned as any of the surveyed applicants “1^st^ preference” factors when determining where a program will fall on their rank list.

**Fig 1.**
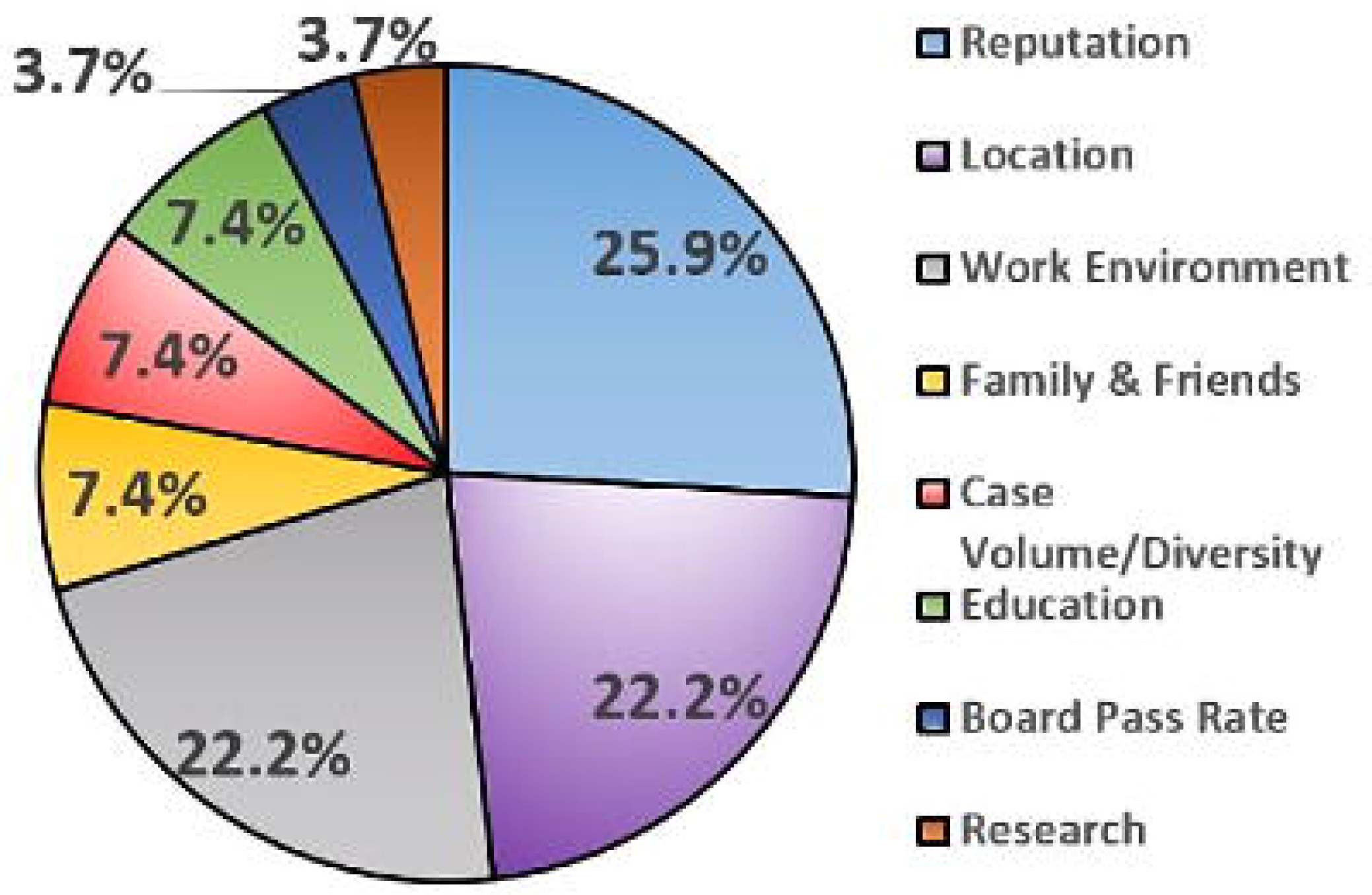

### Factors Considered The 2nd Preference

When considering what factors were the second most mentioned in influencing an applicant’s rank list, approximately 30% of applicants surveyed mentioned that “Work Environment” was their top “2^nd^ preference” pick, as seen in figure 2. This was followed by “Education,” which was the second most common “2^nd^ preference” factor listed at approximately 20%. “Location” and “Reputation” were tied for the thirst most common factors that were mentioned in approximately 13.33% of survey applicants as their “2^nd^ preference” factor. Approximately 6.67% of applicants mentioned that the factor “Fellowships” was their “2^nd^ preference” pick when determining their rank list. The factors “Family & Friends,” “Case Volume/Diversity,” and “Moonlighting” was noted by only 3.33% of applicants as their “2^nd^ preference” item. The factors “Board Pass Rate,” “Research,” “Faculty: Resident Ratio,” “Program Size,” and “Work-Life Balance,” were not listed by any surveyed applicants as being their “2^nd^ preference” factor when determining their rank list.

**Fig 2.**
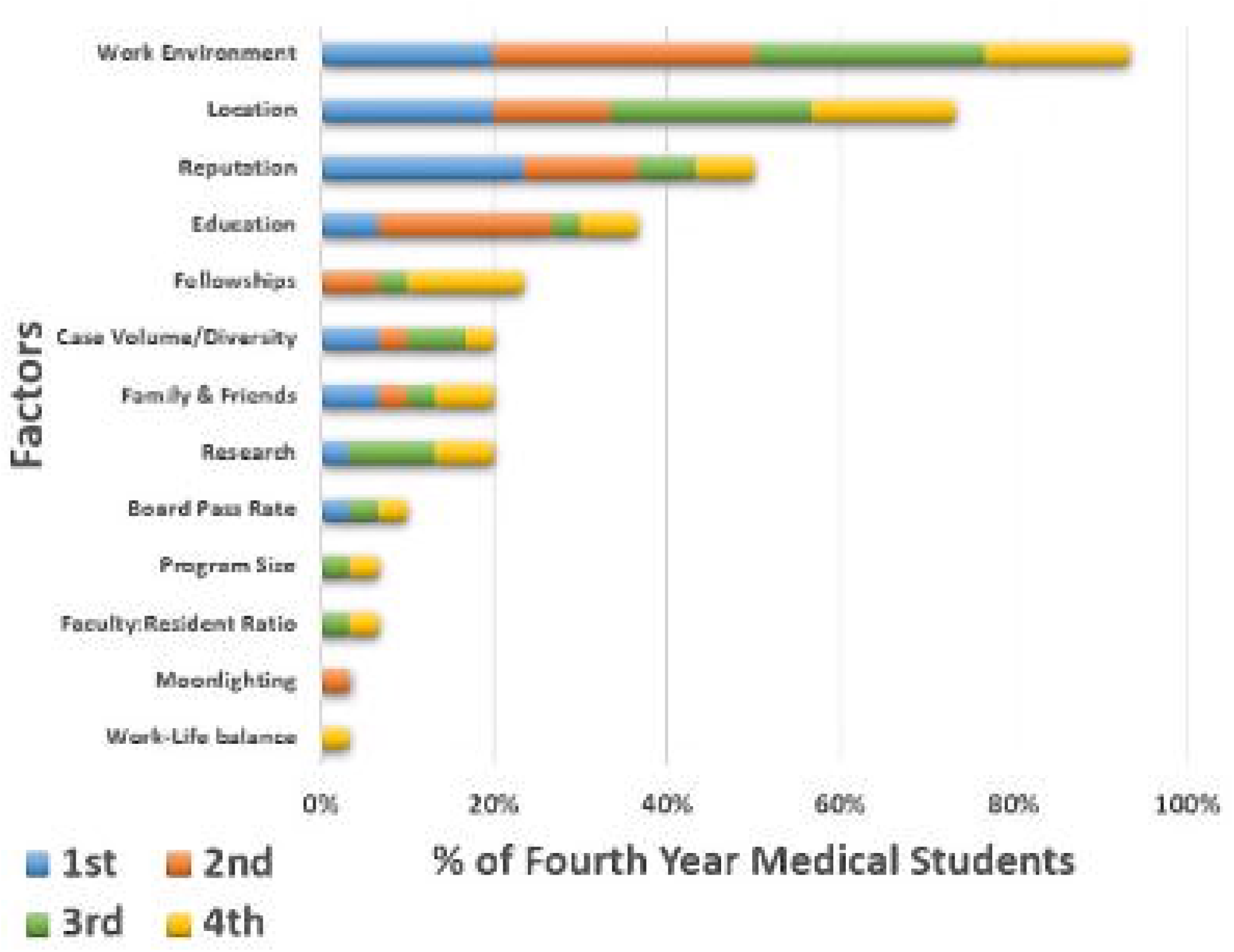

### Factors Considered The 3^rd^ Preference

The most common factor among survey applicants for their “3^rd^ preference” factor in determining their rank list was found to be “Work Environment” yet again, being mentioned in 26.67% of applicant’s responses. In addition to this, the second most common “3^rd^ preference” factor was found to be “Location” at 23.33%, followed by “Case Volume/Diversity which was the third most common “3^rd^ preference” item at 13.3%. The factors “Research” was mentioned in 10% of responses, while “Reputation” appeared in 6.67% of applicants’ responses to their “3^rd^ preference” factor. “Family & Friends,” “Board Pass Rate,” “Fellowships,” “Faculty: Resident Ratio,” and “Program Size” were only mentioned as being the “3^rd^ preference” factor in approximately 3.33% of applicant’s survey responses. The factors “Moonlighting” and “Work-Life Balance” were not mentioned as being any applicant’s “3^rd^ preference” factor in determining the rank list.

### Factors Considered The 4^th^ Preference

The most common “4th preference” factor mentioned in applicant’s surveys were “Location” and “Work Environment,” both being mentioned in approximately 16.67% of responses. This was closely followed by “Fellowships” which was the second most common “4th preference” factor being mentioned in 13.33% of the applicant’s responses. The factors “Reputation,” “Education,” “Family & Friends,” and “Research” was mentioned in only 6.67% of responses. In contrast, the factors “Case Volume/Diversity,” “Board Pass Rate,” “Faculty: Resident Ratio,” “Program Size,” and “Work-Life Balance” were 3.33% of applicant’s “4th preference” response. No applicants listed “Moonlighting” as their “4th preference” response.

## Discussion

Determining what factor’s applicant’s value during the NRMP match process is a simple yet essential piece of information when attempting to fill a residency program with residents that will allow the program to grow and prosper. While the results of this study may not be generalizable to all medical schools due to some of the study limitations encountered, this study does highlight some interesting points that may not be commonly discussed or even thought about. The timing of the survey’s distribution was planned for the day after the match to minimize potential bias answering the survey and to distribute the survey in a time where applicants have developed an idea of what they want out of a program. While the sample size of this study (n = 73) was a limiting factor, we believe the overall design, communication, and timing of the survey’s disbursement is what convinced 30 participants to respond to the survey, giving the survey a response rate of approximately 41%. Considering the complete lack of incentives as well as the time necessary to complete an open-ended survey, having 41% of applicants respond is an accomplishment in addition to a strong point of this study.

### Factors Most Commonly Preferred

The factors that ended up being the most commonly preferred were a program’s reputation at 23.33% of response followed by the location and work environment, which made up 20% of the applicant’s response. While this data is interesting and seems to follow the trends established by previous studies, it can be argued that factors that are out of the program’s control, such as geographic location and national reputation, are not as useful as the factors that a program can change. Work environment is the one factor within the three top factors affecting rank preference that programs have control over. Programs should optimize their work environment, then promote it during recruitment and interview season.

### Factors Most Commonly Mentioned

One trend that highlights the importance of shifting a program’s focus on a superior work environment can be seen in Table 1’s “Mean %” column. The mean percentage column is the aggregate average of the amount of time each factor was listed as a primary, secondary, tertiary, or quaternary preference, allowing us to determine which factors were consistently thought of to be important in the ranking process. Notably, the top three most common “1st preference” factors have had their placements reversed to where the factor “Work Environment” is now shown to be the most mentioned factor overall at 23.35%. Location followed with 18.33% of all submitted answers mentioning the location. In contrast, reputation was only mentioned in 12.5% of all answers despite the survey determining it was applicant’s most common choice for the “1st preference” factor in determining their rank list. The fourth most mentioned factor in the study is “Education” mentioned in 9.18% of all submitted answers, possibly reflecting the idea that applicants value education to a degree, yet believe other factors better define the program’s fit for them.

**Table 1:**
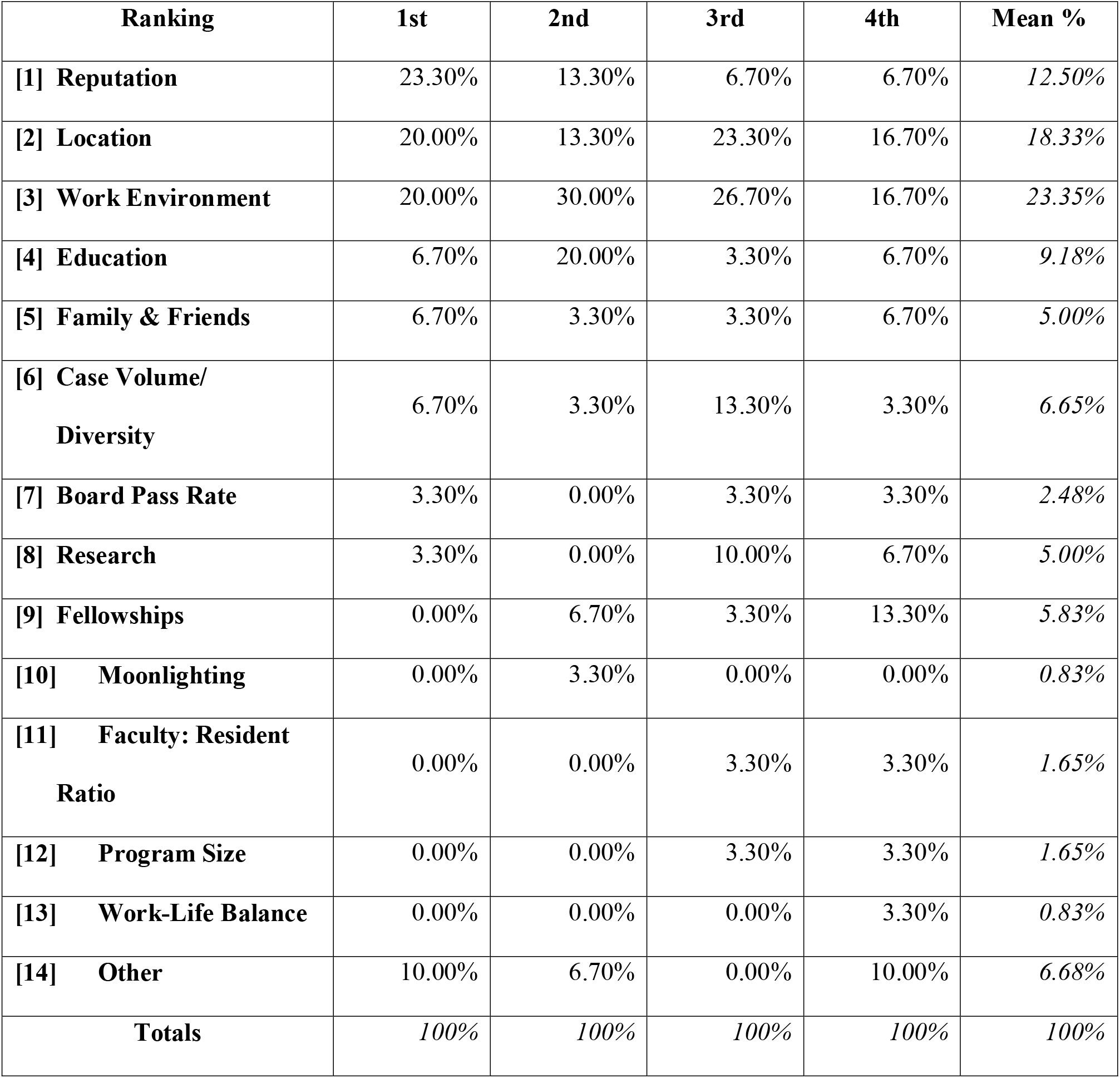
Ranking of Factors Affecting Program Rank List Position Among MS4s.

### Conclusions

Considering approximately 1/4th of all surveyed applicants mentioned that a program’s work environment is within the top four factors they use to rank programs, this further lends to the idea that residency programs should invest in the development and highlighting of a program with a collegial work environment with a team player culture in order to effectively get the highest yield result. While geographic location cannot be changed and national reputation is generally beyond the control of a program immediately, work environment can immediately be optimized by programs to attract top candidates. While these findings seem to endorse previously established trends, it is interesting to see how the factors that are considered the most important by applicants are not necessarily the same factors that are mentioned the most or considered essential when ranking a program. Future studies would need to be considered in order to ensure more definitive and generalizable results.

### Potential Benefits

This study helps identify factors that determine the ranking preferences of future/current radiology applicants, which will help Diagnostic Radiology residency programs better understand the importance of these factors. By acknowledging these ranking preferences, residency programs can work to improve and highlight factors within their control to allow them to match applicants with a higher “fit” by aligning the program’s strengths with the applicant’s preferences.

### Limitations

Possible limitations for this study include many of the same limitations that are seen in survey-based studies such as intrinsic biases due to self-reporting data, possible misinterpretation of questions, evasive answers, and misinterpretation of answers by the investigators. In addition to this, this study did not differentiate applicants based on how competitive they were, which could be an underlying variable that may influence factors applicants find the most important. Because this study only included applicants offered interviews at UTMB as well as only Diagnostic Radiology applicants, this study’s findings hold a bias towards the preferences of Diagnostic Radiology applicants offered interviews at UTMB. With this in mind, results may not be compatible with other specialties or residency programs. Limitations during the development of the survey, such as having clear definitions for “Reputation” or “Work-Life Balance,” may have influenced the categorization of the applicants’ answers. Finally, a small sample size and limited response rate can significantly affect the significance of these studies’ results. Future investigations should be conducted to further refine and build upon the results of this study.

## Data Availability

The authors confirm that the data supporting the findings of this study are available within the article [and/or] its supplementary materials.

